# Nanoparticle capture of urinary lipoarabinomannan for diagnosing childhood tuberculosis

**DOI:** 10.1101/2025.10.13.25337640

**Authors:** Najeeha T. Iqbal, Kumail Ahmed, Brendan T. Mullen, Tania A. Thomas

## Abstract

Urinary assays detecting lipoarabinomannan (LAM) as a diagnostic test for tuberculosis (TB) have limited sensitivity in pediatric populations. We aimed to evaluate a urine processing step using the Ceres TB-Nanotrap to concentrate available LAM antigens and augment LAM detection in urine using the Alere lateral flow assay (LF-LAM). This case-control study recruited children with TB and non-TB controls aged 1-18 years from outpatient clinics. The LF-LAM test was performed before and after concentrating 5mL of urine with 400uL of magnetic TB-Nanotrap particles. Band intensity was measured via visual grading and digital quantification. Urine LAM sensitivity by visual grading was 4.5% (95% Confidence Interval [CI]: 0.3 – 18.5) pre-concentration and 50.0% (95% CI 30.0 – 70.0) post-concentration. Sensitivity by digital quantification was 0.0% (95% CI 0.0 – 15.4) pre-concentration and 63.9% (95% CI 42.8 – 81.4) post-concentration. Specificity was high with both methods (visual grading 90.0% [62.8 – 99.4] pre-concentration, 90.0% [62.8 – 99.4] post-concentration; digital analysis 80.0% [44.4 – 97.5], 90.0% [62.8 – 99.4] respectively). For cases, digital analysis showed an increase in median LF-LAM band intensity from 0.0 arbitrary units (AU) (IQR 0.00 – 32.5) in unconcentrated samples to 98.4 AU (IQR 34.9 – 212.2) in concentrated samples. Concentration with TB-Nanotrap greatly increased sensitivity of urine LAM detection without change in specificity. Digital quantitative analysis further increased sensitivity. Use of TB-Nanotrap and digital quantitative analysis of urine LAM improved the diagnostic accuracy of the LF-LAM assay in this pediatric population and should be validated in larger studies.

## Introduction

To date, an accurate test to confirm a diagnosis of tuberculosis (TB) in children does not exist. The most common method of diagnosing TB relies on detecting mycobacteria from sputum specimens using molecular methods, microscopy, or culture. However, reliance on respiratory specimens does not meet the needs of most children due to logistic and physiologic limitations in collecting a high-quality sample reliably; particularly since childhood TB often represents a paucibacillary disease state. As such, there is a pressing need for novel, pediatric-friendly methods to diagnose TB [1][2]. Evaluating TB biomarkers from urine holds numerous advantages given its relative abundance and the ability to collect it through minimally-invasive means[3][4]. Additionally, a urinary biomarker may obviate the need to localize the exact site of infection, which is especially useful in childhood TB when symptoms may be non-specific and extrapulmonary manifestations are common.

Diagnostic development efforts have focused on detecting lipoarabinomannan (LAM), an outer cell wall mycobacterial glycolipid antigen, from urine specimens among people with active TB disease[5]. Despite progress with commercialized enzyme-linked immunosorbent assays, such as the Alere Determine TB LAM test (LF-LAM), the sensitivity is limited in populations with TB without HIV co-infection[6], including children[7][8]. The LF-LAM assay performs best in people co-infected with TB and advanced HIV (CD4+ T-cells ¡100), with a sensitivity of 56% and specificity of 87% [9]. In addition to concerns about reliability outside of this population, visual assay interpretation of the LF-LAM, the need for CD4 cell count results, and narrow target population have been cited by users as limitations to uptake[10].

Efforts to optimize this assay for expansion into HIV-negative populations include incorporating a urine processing step to concentrate LAM antigens using nanoparticle-based technology to augment detection through commercially available assays. The Nanotrap (Ceres Nanosciences, Manassas VA, USA) employs hydrogel acrylamide microparticles with chemical affinity dyes which attract target analytes in solution. Specimen processing using this technology can concenrate low-abundant molecules and remove proteins or other interfering substances, thereby optimizing the performance of currently available testing platforms. This technology has been applied successfully to infectious diseases such as Lyme, toxoplasmosis, emerging viral pathogens and wastewater analysis[11][12][13]. A TB-Nanotrap has also been created using Reactive Blue 221 as the high-affinity dye that couples well with the 5-methylthio-d-xylofuranose (MTX) epitope of LAM. Initial studies to detect urinary LAM with TB-Nanotraps and the LF-LAM assay among HIV-negative adults with active TB yielded a sensitivity to 96% and specificity of 81% [14]. However, validation studies in pediatric populations are lacking. Therefore, we aimed to evaluate the Ceres TB-Nanotrap as a method of augmenting detection of LAM antigens in urine among children with and without TB using the LF-LAM assay. We also aimed to assess agreement between readers of assay results and the impact of digital LAM assay band quantification on assay performance.

## Methods

### Study Design and Population

This work incorporates a case-control design in a 2:1 ratio among children who were enrolled in a parent study examining a blood-based immunodiagnostic assay. The parent studies recruited children 1-18 years of age from outpatient clinics in the greater Karachi area of Pakistan. Physicians at these clinics referred children who were started on TB treatment based on clinical or microbiologic diagnoses of pulmonary or extrapulmonary TB (“case”). After parental permission and assent were obtained, participants underwent evaluation with standardized data collection including clinical and radiographic information. Specimen collection included blood and urine samples from all, and sputum collection for clinical purposes from those who could spontaneously expectorate for GeneXpert testing. Invasive methods were not pursued to collect respiratory specimens for study purposes. Participants returned after 4-6 weeks for repeat clinical evaluation. Healthy children (“controls”) were recruited from the same settings and enrolled if they had no history of TB disease or TB exposure, no current clinical suspicion of TB, and no history of major illnesses in the last two weeks. After parental permission and assent were obtained, participants underwent evaluation with standardized data collection. “Control” participants returned after 4-6 weeks to ensure clinical stability. Participants (cases and controls) were ineligible as for the parent study if they had received BCG vaccine or tuberculin skin test within the past 8 weeks. This sub-study included participants who had stored urine samples available for additional biomarker testing. Updated NIH consensus criteria were used to further categorize these participants as “confirmed” or “unconfirmed” TB and “non TB controls”[15]. Per these definitions, confirmed cases had at least one sample positive by GeneXpert. Unconfirmed cases had clinical features suggestive of TB and at least two supportive findings between radiographic findings of TB, or immunologic evidence of TB exposure, or clinical response to TB treatment.

### Urine Sample Collection and Processing

All participants provided a spot urine sample, collected non-invasively via clean-catch, which was aliquoted and stored at -80°C for future batch analyses. Urine specimens were thawed and centrifuged at 10,000g for 5 minutes at room temperature. A baseline Alere LF-LAM test was performed using 60µL of the supernatant and interpreted after 25 minutes with use of the Reference Scale Card according to package instructions[16]. The remaining 5mL of the sample was processed with the Ceres magnetic TB-Nanotrap particles. Briefly, 400µL of TB-Nanotrap particles were suspended in 2% SDS solution before adding urine, which was incubated for 30 minutes on an orbital shaker at room temperature. Using a magnetic rack to attract the TB-Nanotrap particles, the supernatant was discarded. Elution buffer (75µL of 2% Tween20 solution) was added and incubated for 15 minutes on an orbital shaker to free LAM from nanocages. Finally, using a magnetic rack to separate TB-Nanotrap particles from the eluted sample, 60µL of the processed sample was removed and loaded on the Alere LF-LAM test strip and interpreted as described above. Visual interpretation: Alere LF-LAM tests were initially interpreted in real time in a non-blinded fashion (KA); images of the test strips were captured using a lightbox with a standardized light source. An independent, blinded reviewer performed a second interpretation of all test results (TAT); any discrepant results were adjudicated by a third independent, blinded reviewer (BTM). All results were recorded qualitatively (positive/negative), as well as quantitatively on a 4-point scale according to band intensity. A positive result required the appearance of any visible band in the “test” window as well as the “control” window. Tests were deemed negative if there was no visible band in the “test” window, but the “control” window showed a visible band.

### Digital Image Analysis

To quantify band intensity in each sample before and after concentration with the TB-Nanotrap, images of each lateral flow assay test strip were photographed from a standard distance using similar lighting conditions. Photos were then imported to ImageJ (National Institutes of Health, Bethesda, MA, USA) for analysis. For each assay image, the peak intensities of the control band and result band were measured. Band intensity was measured in mean arbitrary units. For the purposes of determining performance characteristics, a positive assay was considered one in which the result band had higher intensity than the reference card level 1 band.

### Statistical Analysis

Simple frequencies and measures of central tendency were used to report demographic and clinical data. To explore potential differences in diagnostic accuracy with the TB-Nanotrap, performance measures including sensitivity, specificity, positive predictive value and negative predictive value were calculated as percentages with 95% confidences intervals using confirmed and unconfirmed TB as a composite reference standard. We compared performance measures by visual grading between TB-Nanotrap concentrated and unconcentrated urine. We assessed for incremental change in test characteristics by comparing performance measures by digital analysis between TB-Nanotrap concentrated and unconcentrated urine. Data analysis was performed with SPSS 28.0 (IBM, Armonk, NY).

## RESULTS

### Participant Demographics

We included a total of 32 participants, recruited from April – July 2015 and September 2018 – September 2020, with demographic and clinical data displayed in Table 1. There were 22 participants meeting criteria for the “TB Case” definition which included 10 (31.3%) participants with GeneXpert MTB/RIF confirmed TB and 12 (37.5%) participants who were diagnosed based on clinical criteria. Of the ten participants who were GeneXpert MTB/RIF confirmed, 8 had pulmonary TB. Two had extrapulmonary TB presenting as central nervous system disease with cerebro-spinal fluid samples positive by GeneXpert MTB/RIF. Ten participants were included in the “control” group. Cases (median age = 7.5) were younger than controls (median ages, interquartile range (IQR): 7.5 years (5.0-14.0) and 14.0 years (5.0-18.0), respectively).

**Table 1.** Demographic characteristics. Total of 32 participants, recruited from April – July 2015 and September 2018 – September 2020.

| Characteristic | Overall<br>n = 32 | Cases<br>n = 22 | Controls<br>n = 10 |
| --- | --- | --- | --- |
| Age distribution, n (%) |  |  |  |
| 2-5 years | 9 (28) | 6 (27) | 3 (30) |
| 6-9 years | 7 (22) | 6 (27) | 1 (10) |
| 10-14 years | 7 (22) | 5 (23) | 2 (20) |
| 15-18 years | 9 (28) | 5 (23) | 4 (40) |
| Female, n (%) | 17 (53) | 11 (50) | 6 (60) |
| Cough present, n (%) |  | 19 (86.4) |  |
| Any TB symptom present, n (%) |  | 22 (100) |  |
| Any imaging finding of TB present on<br>chest xray,<br>n (%) |  | 20 (95.2) |  |
| Extrapulmonary TB, n (%) |  | 2 (20) |  |

### LAM Assay Visual Grading

Figure 1 gives an example of the differences in LF-LAM assay results in urine from a single participant with probable TB before and after TB-Nanotrap concentration. The LF-LAM assay was read as negative before concentration and positive after concentration. Figure 2 shows counts of LF-LAM assays by grade for both controls and cases. There were 9 out of 10 control participants who had a negative LF-LAM result before and after TB-Nanotrap concentration. However, one teenage control participant had a positive LF-LAM result that demonstrated a grade 2 band intensity before TB-Nanotrap concentration and grade 1 band intensity after TB-Nanotrap concentration. Among cases, only 1 participant out of 22 had a positive LF-LAM assay with a grade of 1 prior to TB-Nanotrap concentration. After concentration, 11 of 22 had a positive LF-LAM assay, all with a grade of 1. Ages did not differ between cases with positive and negative assays after concentration (p = 0.41). Interobserver agreement was moderate for assay positivity (*κ* = 0.749) and lower for assigning a numeric grade to the assay (*κ* = 0.622). No invalid tests were observed.

**Figure 1.**
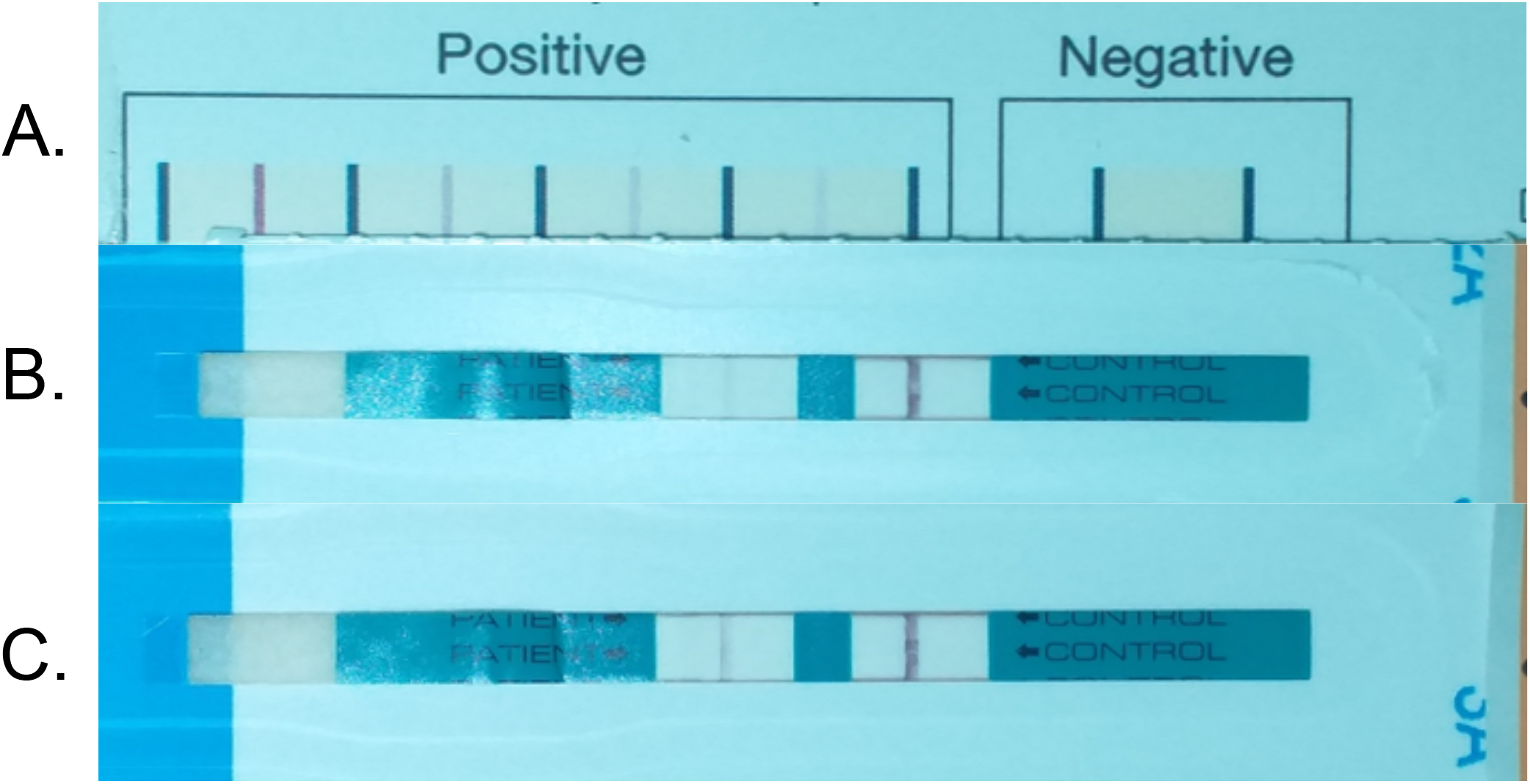
Alere LF-LAM (A) Reference card, (B) assay before TB-Nanotrap concentration, (C) assay after TB-Nanotrap concentration in a participant with probable TB.

**Figure 2.**
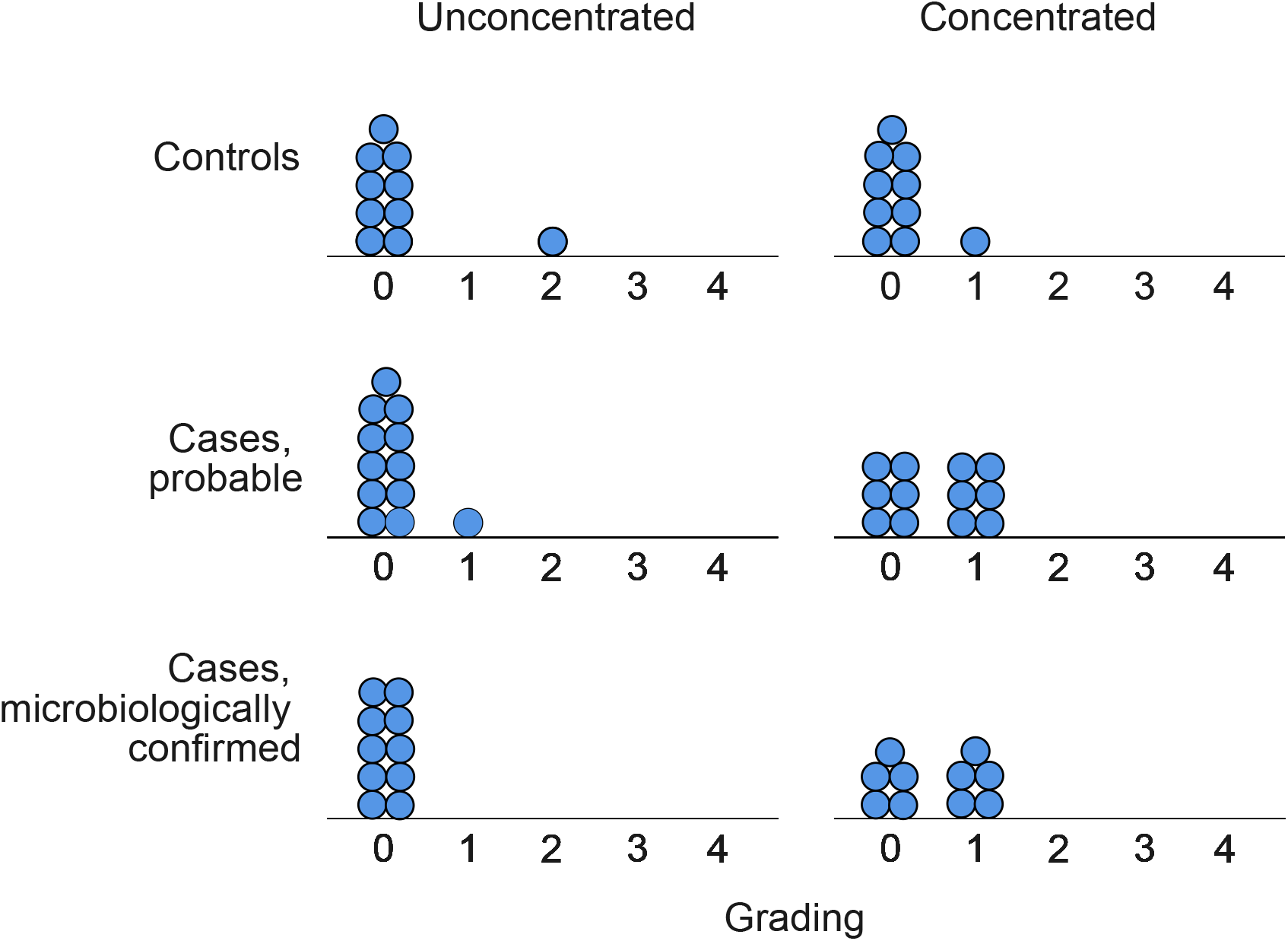
Quantitative changes in band intensity by visual grading after concentration.

### Diagnostic Accuracy of Urine LAM

The diagnostic accuracy of LF-LAM compared to the composite reference standard is shown in Table 2. When compared to the composite reference standard, visual grading of LF-LAM had a sensitivity of 4.5% (95% CI, 0.3 – 18.5) and a specificity of 90.0% (95% CI, 62.8 – 99.4) among cases. After TB-Nanotrap concentration, sensitivity with visual grading increased to 50.0% (95%CI 30.0 – 70.0) with no change in specificity (Table 2). Digital analysis resulted in a further increase in sensitivity of LF-LAM after TB-Nanotrap concentration (sensitivity = 63.%, [95%CI, 42.8 – 81.4]) (Table 2). When stratified by type of case (probable and microbiologically confirmed), sensitivity was similar using both visual and digital analysis. Specificity of the assay with digital analysis was similar across each concentration and analysis method.

**Table 2.** Diagnostic accuracy of unprocessed and TB-Nanotrap concentrated urine LAM by visual and digital analysis.

| Analysis Method<br>and Case Grouping | Sensitivity, % (CI) | Specificity, % (CI) | PPV, % (CI) | NPV, % (CI) |
| --- | --- | --- | --- | --- |
| *Visual Analysis* |  |  |  |  |
| *All Cases* |  |  |  |  |
| LAM without concentration | 4.5 (0.3 – 18.5) | 90.0 (62.8 – 99.4) | 50.0 (3.8 – 96.2) | 30.0 (15.7 – 47.6) |
| LAM with concentration | 50.0 (30.0 – 70.0) | 90.0 (62.8 – 99.4) | 91.7 (68.1 – 99.5) | 45.0 (24.8 – 66.4) |
| *Probable Cases* |  |  |  |  |
| LAM without concentration | 8.3 (0.5 – 31.9) | 90.0 (62.8 – 99.4) | 50.0 (3.8 – 96.2) | 45.0 (24.8 – 66.4) |
| LAM with concentration | 50.0 (23.8 – 76.2) | 90.0 (62.8 – 99.4) | 85.7 (50.6 – 99.1) | 60.0 (35.1 – 81.7) |
| *Microbiologically Confirmed Cases* |  |  |  |  |
| LAM without concentration | 0.0 (0.0 – 30.9) | 90.0 (55.5 – 99.8) | 0.0 (0.0 – 0.0) | 47.4 (42.3 – 52.5) |
| LAM with concentration | 50.0 (18.7 – 81.2) | 90.0 (55.5 – 99.8) | 83.3 (41.3 – 97.3) | 64.3 (48.4 – 77.6) |
| *Digital Analysis* |  |  |  |  |
| *All Cases* |  |  |  |  |
| LAM without concentration | 0.0 (0.0 – 15.4) | 80.0 (44.4 – 97.5) | 0.0 | 26.7 (21.1 – 33.1) |
| LAM with concentration | 63.9 (42.8 – 81.4) | 90.0 (62.8 – 99.4) | 93.3 (73.8 – 99.6) | 52.9 (30.1 – 75.0) |
| *Probable Cases* |  |  |  |  |
| LAM without concentration | 0.0 (0.0 – 26.5) | 80.0 (44.4 – 97.5) | 0.0 (0.0 – 0.0) | 40.0 (32.8 – 47.6) |
| LAM with concentration | 66.7 (34.9 – 90.1) | 90.0 (55.5 – 99.8) | 88.9 (54.4 – 98.2) | 69.2 (49.6 – 83.7) |
| *Microbiologically Confirmed Cases* |  |  |  |  |
| LAM without concentration | 10.0 (0.3 – 44.5) | 80.0 (44.4 – 97.5) | 33.3 (5.1 – 82.4) | 47.1 (38.0 – 56.3) |
| LAM with concentration | 60.0 (26.2 – 87.8) | 90.0 (55.5 – 99.8) | 85.7 (46.6 – 97.6) | 69.2 (50.6 – 83.2) |

Figure 3 shows distribution of assay test band intensity values before and after TB-Nanotrap concentration for controls, probable cases, and microbiologically confirmed cases. Median arbitrary units (AU) threshold for a positive result by visual grading (a level 1 band) was 93.0 AU (IQR 83.5 – 99.0). In controls, median band intensity decreased from 86.0 (0.0 - 194.5) in unconcentrated samples to 3.8 (0.0 – 11.3) in concentrated samples though this change was not statistically significant (*p* = 0.143). In probable cases, median band intensity increased from 8.0 (0.0 - 53.5) to 150.0 (45.3 - 367.0) (*p* = ¡ 0.001). In microbiologically confirmed cases, median band intensity increased from 0.0 (0.0 - 5.5) to 67.0 (26.0 - 122.8) (*p* = 0.011). Among cases, the test band intensity increased after concentration for all with the exception of a single microbiologically confirmed participant. For that child, intensity decreased after concentration from 111.0 to 97.7. In aggregate microbiologically confirmed and probable cases, there was an increase in median band intensity from 0.0 (0.00 – 32.5) to 98.4 AU (34.9 – 212.2) (*p* = ¡ 0.001).

**Figure 3.**
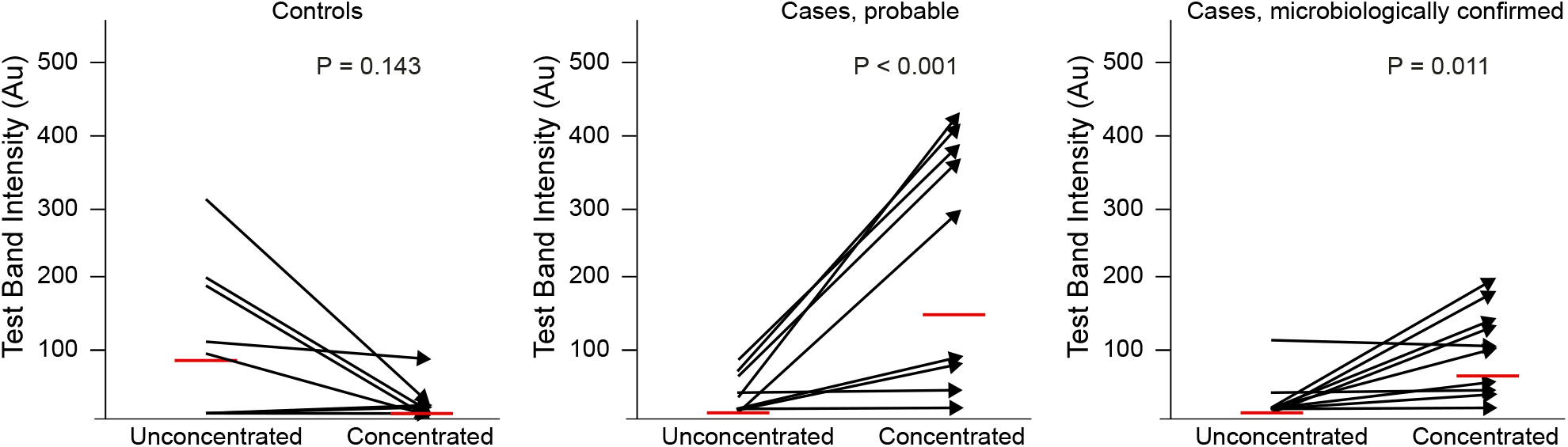
Paired test band intensity values before and after TB-Nanotrap concentration for each control and case. Red line indicates median value

## DISCUSSION

In this cohort analysis of pediatric participants with and without TB, we characterized the diagnostic accuracy of the Alere LF-LAM assay after processing urine samples with the TB-Nanotraps as a means of concentrating any available LAM antigen within the specimen. We found that the TB-Nanotrap increased the overall sensitivity of the urine LF-LAM assay to 50.0% [95%CI 30.0 – 70.0] which approaches LF-LAM sensitivity in patients with advanced HIV[9]. Importantly, concentration increased the sensitivity of LF-LAM from 8.3 [95%CI 0.5 – 31.9] to 50.0 [95%CI 23.8 – 76.2] in participants with probable TB who otherwise did not have a confirmatory sputum test, and there was no difference in detection based on age. Specificity was overall unchanged by the TB-Nanotrap processing step. Urine concentration also resulted in better distinction between the band intensities in both cases and controls, and this was further augmented with the use of a digital reader.

The WHO target product profiles for tuberculosis diagnosis [17] indicate that a non-sputum, low complexity assay should attain a sensitivity of 80%. The use of TB-Nanotrap as presented here does not meet this sensitivity target, but approaches LF-LAM sensitivity in patients with advanced HIV and thus warrants further validation. This study addresses limitations that exist to currently available urine LAM tests, as outlined in the 2019 WHO updates on use of LAM in diagnosis of TB in people living with HIV[18] and in a more recent qualitative survey about use of LAM in TB-endemic regions[10]. Although LF-LAM has a rapid turnaround time and is available as a point-of-care test, the subjective visual assay interpretation, the need for CD4 cell count results, and narrow target population of adults with advanced HIV have been identified as limitations to more widespread uptake. Here, we aimed to address several of these limitations by validating a method of urine LAM concentration with the goal of improving diagnostic accuracy in a pediatric population without HIV. Diagnosis of TB in pediatric populations with presumptive TB remains challenging. TB-Nanotrap worked equally well among children with microbiologically confirmed TB and those with clinically-diagnosed TB. The TB-nanotraps offer a significant additive diagnostic yield, with 50% of clinically diagnosed TB cases having a positive urine LAM after TB-Nanotrap. Our findings suggest that incorporating this processing step to capture urine LAM may be a useful method to increase access to rapid-diagnostic tests for TB in children who otherwise would not have laboratory confirmation of a diagnosis. This is particularly true for children who have negative TB test results by other rapid-diagnostic methods, such as the Xpert MTB/RIF in our cohort. The WHO target product profile report for TB diagnostics identified that a rapid, non-sputum based diagnostic test capable of detecting child TB is of high priority[17]. Urine LAM represents a vital tool for addressing this goal, in that it is rapid and does not require sputum collection. The improved test characteristics shown here with use of nanoparticle concentration warrant further validation in larger cohort studies of patients with and without HIV.

Use of digital quantitative analysis over visual grading further increased TB-Nanotrap concentrated assay sensitivity from 50.0% (95%CI 30.0 – 70.0) to 63.9% (95%CI, 42.8 – 81.4). Our findings of improved test sensitivity with use of digital interpretation of LAM band intensity lend evidence to ongoing developments to implement digital readers as part of urine LAM interpretation[19]. Attempts at utilizing smartphones as an accessible, point of care technology for quantifying assay results have been successful in other lateral flow assays[20][21] and could reasonably be applied to urine LAM as well.

Attempts to quantify the inter-reader variability of LAM assay interpretation have yielded heterogenous results. In 2014 the manufacturer of Alere LF-LAM changed the assay from a 5 point scale to a 4 point scale to assist with discriminating between a positive and negative assay. However, even after this change, studies designed to quantify this variability found significant differences in interpretation at the Grade I cutoff point in the lateral flow LAM assay[22][23]. A recent meta-analysis of LAM assay performance called for consideration of solutions to improve assay interpretation by point-of-care users[24]. Use of objective, quantitative digital grading may be one method of resolving this variability. Several electrochemiluminescence LAM assays are under ongoing investigation to determine if they can provide increased accuracy and precision[25], however protocols for these assays typically required prolonged incubation steps as well as use of a specialized instrument[26]. Methods of processing LAM such as the use of TB-Nanotrap as presented here may have greater potential in maintaining LAM as an instrument-free test in resource limited settings.

The use of quantitative LAM grading may also have implications for clinical outcomes; it has been suggested that an increased quantity of LAM in urine may correlate with more multibacillary disease[14]. In the data presented here, use of TB-Nanotrap significantly increased the quantitative intensity of the assay result band. With higher initial band intensity, the ability to detect quantitative changes in urine LAM over time with treatment could allow for urine LAM to be more rigorously evaluated as a marker of treatment success. All current definitions of TB treatment success rely on sputum and culture-based results[27], highlighting a need for alternative methods of treatment monitoring particularly in children. The cohort presented here was comprised of participants recruited in outpatient and inpatient clinic settings; LAM grading in acutely ill patients with a greater disease burden may be higher at diagnosis. In adults recruited from an inpatient setting, LAM grading has been demonstrated to decrease over time with TB treatment[28]. With improved test precision, reporting LAM as a quantitative numeric grading as opposed to a binary grading may have potential clinical value.

Several limitations exist to the data presented here. Although it is a small cohort, these data serve to address an important gap in data on TB diagnostics between children and adults, as emphasized in the WHO “Roadmap towards ending TB among children and adolescents”[1]. We used a composite clinical reference standard rather a purely microbiological reference standard, which may have impacted the performance characteristics by including participants with more paucibacillary disease. However, this composite reference standard more accurately reflects the high proportion of children who are diagnosed with TB without microbiologic confirmation[29] and adheres to consensus guidelines for studies on TB diagnostics. Other limitations include the TB-Nanotrap methodology as it relates to the goal of urine LAM being accessible as a bedside test. As it stands, methods employed here require laboratory space, basic equipment, personnel, and time that may limit clinical implementation. These initial data support the continued refinement of the TB-Nanotrap concentration methodology to ensure its accessibility, given the improvements seen in LAM test characteristics. TB-Nanotrap concentration here did not significantly improve the specificity of the LAM assay. Several explanations have been proposed for limitations to specificity for urine LAM. In children, urine collection bags may be prone to becoming contaminated with commensal skin bacteria that might cross-react with LAM assay antibodies[30]. Whether through disseminated infection or contamination, nontuberculous mycobacteria have been shown to cross react with urinary LAM assays [31]. It has also been shown that LAM assays can be falsely positive in the presence of other skin and oral flora including Nocaria spp., Actinomycetes spp., and Candida albicans [32]. Efforts are underway exploring other antibody combinations to target MTB urine LAM more specifically [26].

Overall, data presented here on diagnostic accuracy of urine LAM with nanoparticle concentration and digital analysis demonstrate improved accuracy in a pediatric population. In addition to further validation studies in larger pediatric cohorts, future studies should be done to evaluate the performance of urine LAM with nanoparticle concentration in broader patient populations with limited TB diagnostic tools, and the impact of its use on clinical outcomes.

## Data Availability

All data produced in the present study are available upon reasonable request to the authors

## ETHICS APPROVAL

This protocol was approved by the Ethical Review Committee at Aga Khan University (AKU ERC# 2924 and 019-0294-2363), the Institutional Review Board of Civil Hospital Karachi & Dow University of Health Sciences, Karachi (IRB #450), and the Institutional Review Board of University of Virginia (IRB-HSR# 17239). Parents/caregivers of eligible children underwent the informed consent process and provided written parental permission for their child’s participation.

## FUNDING

This work was funded by the University of Virginia Global Infectious Disease Institute and the National Research Program for Universities (NRPU), supported by Higher Education Commission (HEC) Pakistan. Award No: 5213/Sindh/NRPU /R&D/HEC/2026. This work was also supported by NIH T32 AI007044.

## CONFLICTS OF INTEREST

The authors declare no conflicts of interest

## Notes

### Competing Interest Statement

The authors have declared no competing interest.

### Author Declarations

Ethical Review Committee at Aga Khan University gave ethical approval for this work (AKU ERC# 2924 and 019-0294-2363) The Institutional Review Board of Civil Hospital Karachi & Dow University of Health Sciences Karachi (IRB #450) gave ethical approval for this work The Institutional Review Board of University of Virginia (IRB-HSR# 17239) gave ethical approval for this work. Parents/caregivers of eligible children underwent the informed consent process and provided written parental permission for participation.

